# Optimized Surveillance Testing for Toxigenic *Clostridioides difficile* to Scale Prevention Programs Across Clinical Settings

**DOI:** 10.64898/2026.07.22.26358596

**Authors:** LM Cersosimo, N Correa, ML Delaney, L DelloStritto, M Misialek, M Klompas, M Baker, L Bry

## Abstract

*Clostridioides difficile* is the leading cause of healthcare acquired infections (HAIs). Healthcare systems lack scalable strategies to identify patients colonized with toxigenic strains to inform prevention programs. We developed a scalable program using rectal swabs, collected for vancomycin resistance Enterococci surveillance, with *ex vivo* amplification of *C. difficile* in Cdiff Banana Broth (BB). Optimized confirmatory testing of BB-positive tubes used the rapid *C. difficile* QuickCheck assay with Cepheid’s Xpert® C. difficile/Epi (Xpert) to confirm toxin gene carriage for QuickCheck results of *C. difficile* glutamate dehydrogenase-positive (GDH) and antigenic toxin-negative. BB with confirmation demonstrated 92% specificity and 100% sensitivity, to a limit of <5 colony forming units (CFU)/swab, versus published values of 100/swab by ChromID® *C. difficile* agar and 460/swab by direct Xpert swab testing. The two-step confirmation detected toxigenic *C. difficile* in cases missed by other methods.

Prospective testing of 797 VRE swabs from 538 ICU and inpatient oncology patients seen at an academic medical center and two community hospitals demonstrated 11% positivity for toxigenic and 4% for non-toxigenic C. difficile. Among swabs, 32% of toxigenic *C. difficile* were resulted at 24 hours and 68% at 48 hours. Addition of 17% glycerol to BB-positive aliquots, for –80°C storage, supported 100% retrieval of *C. difficile* 7 months later. BB with confirmation provides a more sensitive method to detect patient colonization with toxigenic C. difficile and, by using VRE surveillance swabs, does not require additional patient samples. The simplified approach can be deployed across healthcare settings to support *C. difficile*-prevention programs.

**Importance:** *C. difficile* infections are the most prevalent HAIs, causing substantive mortality, morbidity, and economic burden. Among HAIs, *C. difficile* remains the one pathogen lacking scalable surveillance strategies for clinical microbiology labs. We present an optimized strategy to enable *C*. *difficile* surveillance testing across broad clinical settings to enable prevention programs for this pathogen.

## Introduction

*Clostridioides difficile,* the cause of pseudomembranous colitis, is a toxigenic, spore-forming, obligate anaerobe. CDI is frequently triggered with antibiotic exposures that ablate protective commensals (1, 2). Patient colonization with toxigenic strains is a dominant risk factor for CDI, with relative risks of 9.3 to >20 identified in ICU patients (1, 3). Asymptomatic carriers are also dominant reservoirs for strain transmission through shed spores (1, 4, 5).

C. *difficile* infections (CDI) account for 12% of healthcare-associated infections (6), with an estimated 500,000 cases per year and economic burden in the US that exceeds $2 billion/year (7). With 35.7 million hospital admissions/year in the US, for an estimated 28-30 million individuals (8), CD-colonization rates of 10%, to more than 20% of hospitalized patients, suggest that 2.8 to more than 5 million patients/year may unknowingly be bringing in toxigenic CD strains during their hospital stays, contributing to the high disease burden and transmission of CD within nosocomial settings.

The lack of *C. difficile* surveillance programs thus contributes to the high disease burden seen in the US and abroad (1, 9). Surveillance programs have utilized combinations of selective anaerobic culture, commercial enzyme immunoassays (EIAs), and PCR to detect toxigenic *C*. *difficile*. Culture-based techniques include alcohol shock of primary samples to select for spores and use of selective media including cefoxitin-cycloserine-fructose agar (CCFA) or broth, and ChromID® C. difficile agar (1, 4, 10, 11). However, as these methods require specialized resources including anaerobic chambers and personnel with expertise in handling anaerobes (12), they are not accessible to most clinical laboratories and present barriers for establishing *C*. *difficile* surveillance programs. Non-culture-based methods include direct testing of patient samples with enzyme immunoassays (EIAs) to detect *C*. *difficile’s* glutamate dehydrogenase (GDH) and toxins, and PCR-based testing for these targets which have been used to support CD surveillance in highly vulnerable patient populations (13, 14). However, existing CLIA-approved tests are optimized for the higher pathogen burdens and toxin levels that occur in symptomatic patients and can miss low levels of colonization, including cases where toxin is not expressed (4). These approaches can also present limitations for throughput and costs within a clinical lab setting.

To reduce complexities for implementing scalable CD surveillance programs, our group and others have used rectal swabs collected for detection of vancomycin resistant *Enterococci* (VRE) to also support *C. difficile* surveillance, initially by plating swabs to ChromID® *C. difficile* agar (1, 4, 5, 15). However, the need for anaerobic equipment and expertise limited broader adoption of this methodology. To address these limitations, we leveraged selective C. diff Banana Broth (BB)(16), a *C. difficile* selective and enrichment broth media originally developed for environmental recovery of *C*. *difficile*. The media comes in screw-capped tubes that maintain anaerobic conditions once sealed. The formulation (Table 1) includes thioglycolic acid, L-cystine, lysozyme, mannitol, and sodium taurocholate to promote *C*. *difficile* germination and growth, and cefoxitin and cycloserine to inhibit other species (16). For environmental testing, pre-moistened swabs are passed over surfaces, placed in the BB tube and incubated aerobically for up to 72h. Samples positive for *C. difficile* growth change from red to yellow and are further tested with confirmatory methods (16–18).

**Table 1:** Banana broth formulation from Cadnum, et al.

| Ingredient | Quantity/liter |
| --- | --- |
| Brucella Broth | 28.0g |
| Vitamin K1 solution (1mg/ml) | 1.0ml |
| Hemin solution (5mg/ml) | 1.0ml |
| Sodium Bicarbonate solution (20mg/ml) | 5.0ml |
| D-Mannitol | 6.0g |
| Neutral Red solution (1%) | 5.0ml |
| Sodium Taurocholate | 0.5g |
| Lysozyme | 5.0mg |
| D-Cycloserine | 500.0mg |
| Cefoxitin | 16.0mg |
| Agar | 1.0g |
| Thioglycolic Acid (mercaptoacetic acid) | 1.0g |
| L-Cystine | 1.0g |

Legend: Ingredients for the preparation of Banana Broth from Cadnum, et al. (16)

We validated BB’s performance for *C. difficile* detection from VRE rectal swabs, using rapid antigenic testing to confirm production of *C. difficile* glutamate dehydrogenase (GDH) and toxin, followed by Xpert® C. difficile/Epi for toxin gene carriage in cases of BB+ tubes that were GDH+ and negative for antigenic toxin, a situation that occurs when *C. difficile* populations are not sufficiently stressed to induce express their toxin genes. Analyses determined BB’s limit of detection (LoD), sensitivity, specificity, and isolate recovery to support strain-level analyses. Our approach overcomes major barriers to *C. difficile* surveillance, including leveraging existing VRE rectal swabs, removing the need for dedicated resources and expertise in anaerobic cultivation, and using *C. difficile* confirmatory methods that are routinely available in Clinical Laboratories.

## Materials and Methods

### Bacterial strains (Table 2)

*C. difficile* quality control (QC) strains used ATCC 43255 (genomic clade 1), epidemic strain UK1 (clade 2, binary toxin positive), E15 (clade 3), strain CD02 (clade 4), and strain CD55 (clade 5). The negative QC strain used *Clostridium perfringens* ATCC 13124.

**Table 2:** Banana Broth (BB) results with non-*C. difficile* species.

| Microbial Species | Strain | BB 24h | BB 48h |
| --- | --- | --- | --- |
| *Bacteroides ovatus* | ATCC 8483 | Negative | Negative |
| *Bacteroides thetaiotaomicron* | Isolated from BB | Negative | Negative |
| *Candida albicans* | Isolated from BB | Negative | Negative |
| *Clostridium acetobutylicum* | ATCC 824 | Negative | Negative |
| *Clostridium baratii* | Isolated from BB | Positive | Positive |
| *Clostridium butyricum* | MIYAIRI 588 | Positive | Positive |
| *Clostridium butyricum* | Isolated from BB | Positive | Positive |
| *Clostridium clostridioforme* | Clinical | Negative | Negative |
| *Clostridium indolis* | Clinical | Negative | Negative |
| *Clostridium innocuum* | Clinical | Negative | Negative |
| *Clostridium innocuum* | Isolated from BB | Positive | Positive |
| *Clostridium perfringens* | ATCC 13124 | Negative | Negative |
| *Clostridium perfringens* | Isolated from BB | Positive | Positive |
| *Clostridium ramosum* | Clinical | Negative | Negative |
| *Clostridium ramosum* | DSM 1402 | Negative | Negative |
| *Clostridium sardiniense* | DSM 599 | Positive | Positive |
| *Clostridium scatologenes* | ATCC 757 | Positive | Positive |
| *Clostridium scindens* | ATCC 37504 | Positive | Positive |
| *Clostridium tertium* | Clinical | Negative | Negative |
| *Enterococcus faecalis* | Isolated from BB | Negative | Negative |
| *Enterococcus faecium* | Isolated from BB | Negative | Negative |
| *Escherichia coli* | Isolated from BB | Negative | Negative |
| *Morganella morganii* | Isolated from BB | Negative | Negative |
| *Paraclostridium bifermentans* | ATCC 638 | Positive | Positive |
| *Paraclostridium bifermentans* | Isolated from BB | Positive | Positive |
| *Phocaeicola vulgatus* | Isolated from BB | Negative | Positive |
| *Proteus mirabilis* | ATCC 29906 | Negative | Negative |
| *Staphylococcus aureus* | ATCC 25923 | Negative | Negative |

Legend: BB results at 24 and 48h of incubation, as described in the methods, when inoculated with defined species.

BB specificity studies used the following clinical and type strains: *Bacteroides ovatus* ATCC 8483, *Clostridium acetobutylicum* ATCC 824, *Clostridium perfringens* ATCC 13124, *Clostridium butyricum* MIYAIRI 588, *Clostridium scindens* ATCC 37504, *Clostridium sardiniense* DSM 599, *Clostridium scatologenes* ATCC 757, *Paraclostridium bifermentans* ATCC 638, *Proteus mirabilis* ATCC 29906, and *Staphylococcus aureus* ATCC 25923. Clinical isolates included *Bacteroides thetaiotaomicron*, *Candida albicans, Clostridium baratii, Clostridium butyricum, Clostridium clostridioforme, Clostridium indolis, Clostridium innocuum, Clostridium perfringens, Clostridium ramosum, Clostridium tertium, Enterococcus faecalis*, *Enterococcus faecium*, *Escherichia coli*, *Morganella morganii, Paraclostridium bifermentans,* and *Phocaeicola vulgatus*.

### Ethical oversight and sample collection

Assay validation and prospective surveillance were conducted from May to November 2024 under IRBs 2005P001742 (Mass General Brigham, Bry PI, for assay validation and testing at BWH, FH and NWH) and 2022P000819 (Mass General Brigham, Baker PI, for use of results in prospective surveillance). Rectal swabs, submitted for VRE ICU and inpatient oncology unit surveillance at Brigham and Women’s Hospital (BWH), Brigham and Women’s Faulkner Hospital (FH) and Newton Wellesley Hospital (NWH) were retrieved after completion of VRE surveillance testing using the Crimson system (1, 19). Swabs were stored after VRE testing at 4°C for 2 hours to 3 days prior to BB testing which were shown to preserve spore viablity for detection (1). Prospective *C. difficile* surveillance results from VRE swabs submitted to the BWH Clinical Microbiology laboratory were reported to the infection control teams at BWH in support of clinical trial NCT05389904 (Figure 1B).

**Figure 1.**
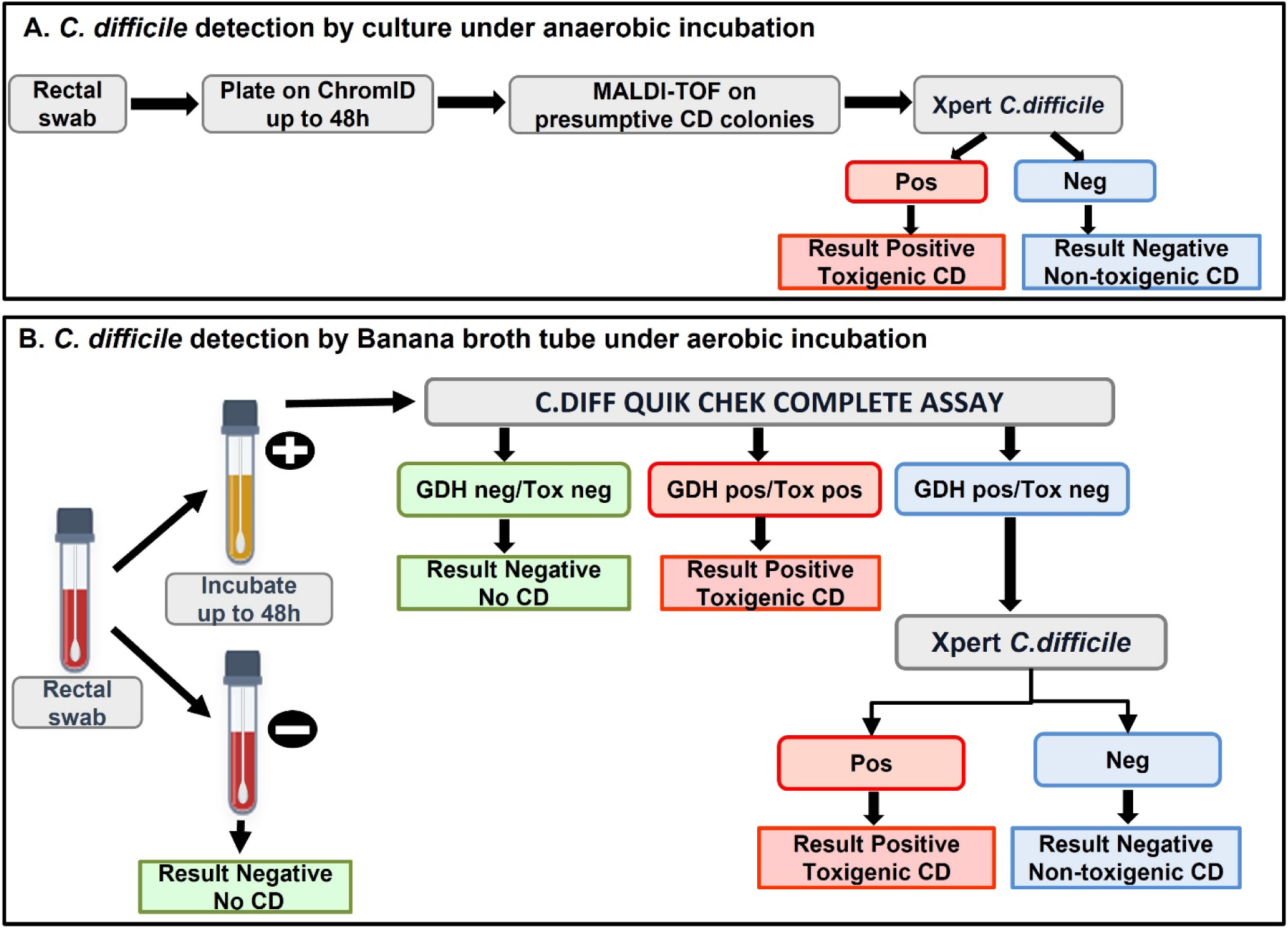
*C. difficile* surveillance testing workflows: **Legend**: A. Workflow using ChromID plates under anaerobic incubation. Identification of CD by MALDI-TOF and testing of CD toxin by Cepheid Xpert (PCR). B. Optimized workflow using BB as screening method without the need for onsite anaerobic chamber. BB-positive tubes are tested by rapid C.diff complete EIA. Samples that are GDH positive and Tox negative undergo Xpert for a final result of positive or negative for toxigenic CD

### Banana Broth cultivation for *C. difficile*

Within a biosafety cabinet, each rectal swab was inserted into BB tubes (Hardy Diagnostics, Santa Maria, CA). The top of the swab stem was snapped off to allow the swab to fit, and the tube cap replaced and tightly closed. Sealed tubes were incubated in a 37°C aerobic incubator and checked at 24h and 48h for turbidity and color change from red to yellow. At 24h of incubation, tubes showing any yellow coloration from the tube base were evaluated for possible *C. difficile*. All other tubes were incubated to 48h for final review and resulting. Tubes showing no color change by 48h of incubation were resulted as negative. Quality Control testing of each lot of BB used *C. difficile* ATCC 43255 as the positive control and *Clostridium perfringens* ATCC 13124 as the negative control.

### ChromID® *C. difficile* agar

(ChromID®, bioMérieux, Marcy-Etoile, France): VRE rectal swabs were struck to ChromID followed by anaerobic incubation in a Coy chamber at 37°C with an atmosphere of 10% hydrogen, 10% carbon dioxide, and 80% nitrogen (20). For ChromID evaluation, plates were examined at 24 and 48h for grey to black pigmented *C*. *difficile* colonies which were confirmed by VITEK-MS (bioMérieux, Marcy-Etoile, France) and tested for the presence of the *tcdB* gene (Toxin B) using the Xpert® C. difficile/Epi kit (Cepheid, Sunnyvale, CA) (Figure 1A). Prior studies by Worley et al. indicated a limit of detection for *C. difficile* at 100 CFU/swab with this method (1).

### C.diff Quik-Chek Complete assay

(CDQCC, TechLab, Blacksburg, VA): is an enzyme immunoassay (EIA) that detects *C*. *difficile* GDH and toxins A and B. This test was performed following the manufacturer’s instructions for liquid stool samples by collecting 100 uL of positive (21)BB from areas with yellow color, commonly at the bottom of the tube, and adding the material to 650 uL of diluent. Samples negative for GDH and toxin were reported as negative, whereas those positive for both GDH and toxins were reported as positive for toxigenic *C. difficile*. Samples that were positive for GDH and negative for toxin were further tested for toxin gene carriage using the Xpert® C. difficile/Epi.

### Xpert C. difficile/Epi

(Xpert, Cepheid, Sunnyvale, USA): detects the toxin B gene and presumptive identification of 027/NAP1/BI strains by detection of binary toxins and a deletion in the negative regulator of toxin production. Cepheid’s published limit of detection for toxigenic *C. difficile* on primary samples, at 95% sensitivity, for Xpert® *C. difficile/*Epi, is 460/sample (21, 22). Xpert® *C. difficile/*Epi was performed by dipping a sterile swab in the positive BB tube and mixing with the kit’s buffer. The inoculated buffer was then transferred to the cartridge and placed in the GeneXpert Infinity system.

### BB Limit of Detection

A spore stock of *C*. *difficile* ATCC 43255 at 1×10^6^ spores/ml was serially diluted to 1×10^1^-10^5^ spores/ml, followed by transfer of 100uL from each dilution to a 5ml polystyrene round bottom tube, for approximately 1 to 10^4^ spores per inoculum. A BBL CultureSwab (Becton, Dickinson and Company, Franklin Lakes, NJ) was added to the tube to absorb the solution and placed into a BB tube and incubated in a regular incubator at 37°C. Tubes were checked at 24 and 48h for turbidity and color changes. Analyses evaluated four swabs per input spore concentration.

### BB Cross-reactivity

To assess the specificity of BB for *C. difficile,* overnight cultures of the species listed under the section “Bacterial strains” were grown in pre-reduced BHIS (Anaerobe Systems). Swabs were dipped into the cultures, in triplicate, and inoculated into BB tubes. BB tubes were evaluated at 24 and 48 hours for development of a yellow color. Tubes turning yellow were plated to pre-reduced Brucella agar to confirm growth.

### Quality Control Program

Quality control testing was performed for each lot or shipment of testing reagents, as well as monthly for BB, ChromID, CDQCC, and Xpert® *C. difficile/*Epi. BB and ChromID were evaluated with *C. difficile* ATCC 43255 as a positive control and *C. perfringens* ATCC 13124 as a negative control. Preparations of 10^4^ spores/mL, as noted under “BB Limit of Detection,” were also used to support BB QC testing to limit needs for onsite anaerobic cultivation. Controls provided in the CDQCC test kit were tested following the manufacturer’s instructions. For Xpert® *C. difficile/*Epi QC, a molecular *C. difficile* control set containing positive and negative matrices was used (M108, Quidel Corporation, Athens, OH).

### *C. difficile* strain retrieval from BB positive tubes

To evaluate the preservation of *C. difficile* strains in BB tubes, for support of strain-level analyses at a later date, aliquots from seven *C. difficile-*positive BB tubes and one inoculated with *C*. *difficile* ATCC 43255 were each added to 0.4 ml of 60% glycerol (final concentration of 17% glycerol) for storage at –80°C. *C. difficile* recovery was evaluated in samples prior to freezing and at weeks, 1, 4, 7, and 12 months after freezing. Strain retrieval used ChromID agar inoculated with 50 µl of frozen, glycerol-preserved material followed by anaerobic incubation for 24-48h in a Coy anaerobic chamber. Colonies identified as presumptive positive *C*. *difficile* were confirmed by Vitek MS.

### DNA Extraction from BB for Nucleic Acid Analyses

To evaluate BB’s performance in detecting diverse *C. difficile* strains, representative strains from *C*. *difficile* genomic clades 1 to 5, with concentrations from 10^1^-10^3^ CFU/ml, were added to BB and incubated for 48h. From positive BB tubes, a one ml aliquot of each sample was centrifuged at 5,000 x g for 10 min. DNA was extracted from the pellet using the EZ1 & 2 DNA Tissue Kit (Qiagen, Germantown, MD). A total of 180 µl of G2 buffer was added to the pellet, vortexed, and 20 µl lysozyme (50 mg/ml) added. Tubes were incubated at 37°C for 1h for cell lysis and extracted with the EZ1 liquid handler using an elution volume of 50 µl. DNA concentrations were measured via Qubit 3.0 (Thermo Scientific, Waltham, MA) and found to be at ≤5ng/uL from the frozen materials, and higher from re-cultivated strains. The *tcdB* (*C*.*difficile* toxin B) and *gdh* (glutamate dehydrogenase) genes were targeted via qPCR in a Quant Studio 12 Flex instrument (Thermo Scientific, Waltham, MA) with the PowerUp SYBR Green Master Mix (Applied Biosystems, Foster City, CA) and primers tcdB-F (5’-CTGGAGAATGGAAGGTGGTT-3’) and tcdB-R (5’-TTGATGGTGCTGAAAAGAAGTG-3’) and gluD-F (5’-ATGCAGTAGGGCCAACAAAA-3’) and gluD-R (5’-TTCCACCTTTACCTCCACCA-3’). qPCR was performed on each target in triplicate for each sample, negative control, and positive control (*C*. *difficile* ATCC 43255 DNA) under the Master Mix manufacturer’s suggested conditions: Hold Stage (50°C 2 min, 95°C 2 min), PCR Stage (95°C 15s, 60°C 1 min), and Melt Curve (95°C 15s, 60°C 1 min, 95°C 15s). All BB-frozen samples of toxigenic *C. difficile* demonstrated positive signal for both targets.

### Statistical analyses

Sensitivity, specificity and 95% confidence intervals (CI) were calculated in GraphPad Prism version 11.0.0 using the Wilson-Brown method (GraphPad Software, San Diego, California). Comparisons evaluated the performance of BB with confirmation versus *C. diff* ChromID agar or versus direct swab testing by Xpert® C. difficile/Epi.

## Results

We evaluated BB’s performance to support detection of toxigenic *C. difficile* from VRE rectal swabs. Limit of detection studies inoculated BB tubes with 10^0^-10^4^ spores/ml of *C. difficile* ATCC 43255. By 48h, 75% (3 of 4 replicates) of the tubes inoculated with approximately 1 spore/ml turned yellow while 100% of tubes inoculated with 10^1^ or higher inocula turned yellow by 24-48h. BB’s limit of detection was estimated at <5 spores by 48h of incubation (Figure 2). BB tubes were also inoculated with *C. difficile* strains belonging to genomic clades 1 through 5 (Methods), at different concentrations (10^1^ to 10^3^ CFU/ml) to evaluate bacterial growth and presence of inhibitors for PCR-based confirmatory methods such as Xpert. All clades, at the three concentrations, turned BB positive by 24-48h. Xpert results confirmed the presence of toxigenic *C. difficile* for all clades including the binary toxin-positive strain UK1.

**Figure 2.**
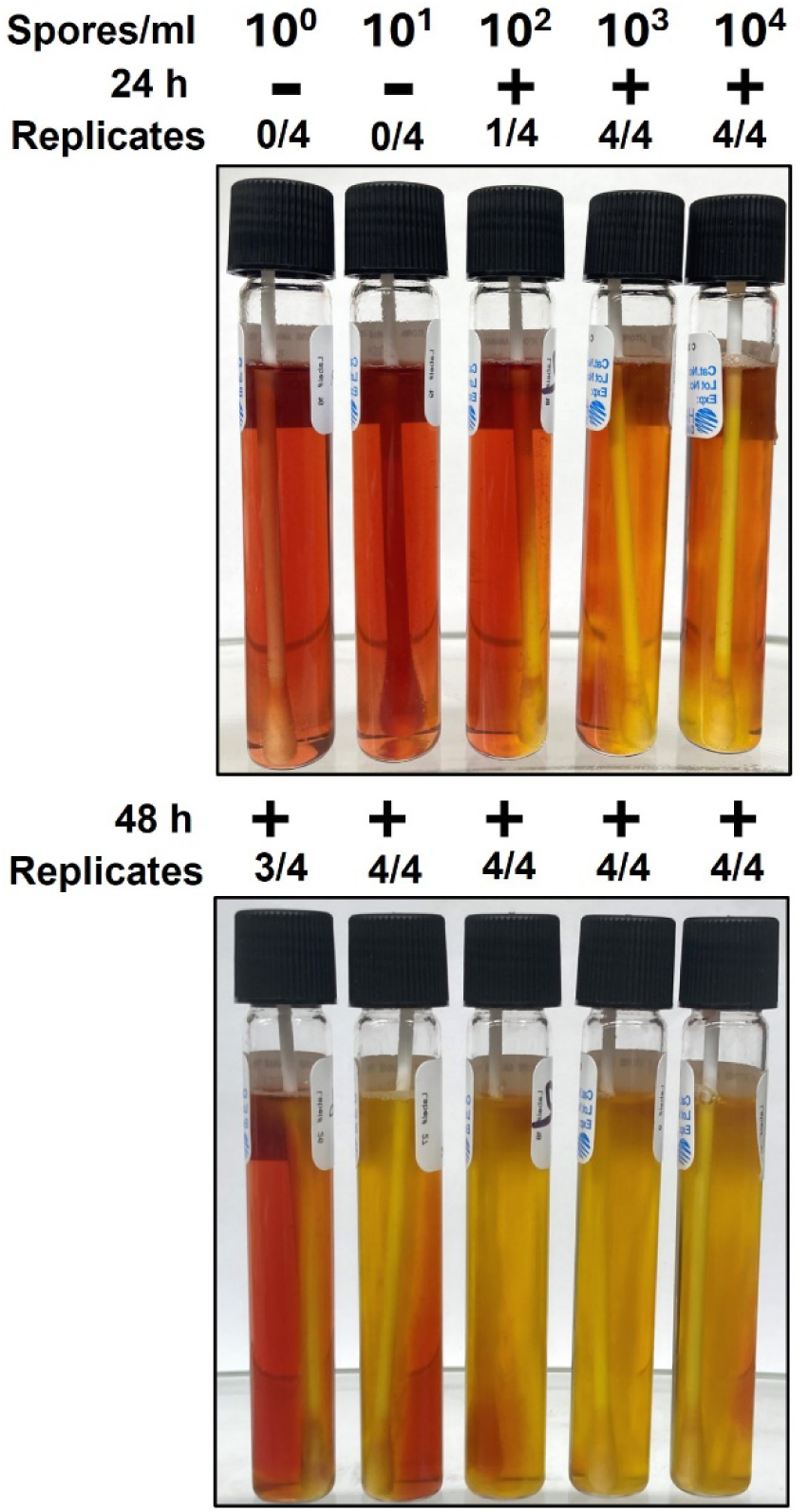
BB Limit of Detection: **Legend:** Photographs of BB tubes inoculated with dilutions of *C. difficile* strain ATCC 43255 from 100 to 104 spores and read at 24h and 48h. A change of color from red to yellow indicates a positive reaction. Limit of detection was estimated at <5 spores by 48h of incubation.

### BB sensitivity and specificity as compared with ChromID culture

Fifty VRE swabs tested by ChromID agar culture and Xpert confirmation (20 positive, 30 negative) were evaluated with BB. Compared with ChromID culture with Xpert confirmation (Figure 1A), BB with confirmation for toxigenic *C. difficile* by Xpert demonstrated a sensitivity of 100% and specificity of 97% by 48h (95%CI 0.83 to 1.00). To further evaluate BB’s performance in prospective surveillance relative to ChromID/Xpert, 100 prospectively collected VRE swabs were tested. Presumptive *C. difficile* colonies from ChromID and BB positive samples were confirmed as toxigenic *C. difficile* by Xpert. ChromID/Xpert identified 5 samples with toxigenic *C. difficile* while the BB identified these 5 samples and an additional 2 samples, illustrating capacity for BB’s *ex vivo* enrichment to enhance detection of low burdens of colonization with toxigenic C. *difficile*. The sensitivity and specificity for BB with Xpert confirmation was 100% and 98% respectively. BB-positive tubes that tested negative for toxigenic *C. difficile* included those carrying non-toxigenic *C. difficile* strains or commensal species able to ferment the mannitol and grow in the presence of the media’s antibiotics, including *Phocaeicola vulgatus* and other commensal *Clostridia.* The other commensal species more often turned the media positive by 48h of incubation (Table 2).

### BB/CDQCC/Xpert

To streamline confirmatory testing and identify a cost-effective methodology using existing FDA-approved clinical testing for *C. difficile*, we evaluated BB+ tubes with the rapid C. diff Quik Chek Complete EIA (CDQCC), which detects antigenic production of *C. difficile’s* glutamate dehydrogenase (GDH) and toxins A/B. For cases where CDQCC identified presence of *C. difficile,* per the positive GDH reaction, but not antigenic toxin expression, samples proceeded to Xpert confirmation of toxin gene carriage (Figure 1B). Forty VRE rectal swabs were prospectively processed by both ChromID agar and BB. BB-positive samples underwent confirmatory testing by CDQCC, followed by Xpert on samples that were GDH positive and negative for toxin A/B. *C. difficile* presumptive colonies isolated from ChromID were confirmed by Xpert.

BB testing with CDQCC and Xpert confirmation when needed, was positive for toxigenic *C. difficile* in 12.5% of samples (5/40) and negative in 87.5% (35/40). In contrast, ChromID/Xpert identified 5% (2/40) of samples as toxigenic *C. difficile*-positive and 95% (38/40) as negative. BB with confirmation detected the 2 swabs also positive by ChromID, and 3 additional swabs missed by ChromID. Sensitivity and specificity of the BB/CDQCC/Xpert to confirm the presence of toxigenic *C*. *difficile* is 100% and 92%, respectively (Table 3), and demonstrated enhanced sensitivity for detecting *C. difficile* colonization on rectal swabs versus ChromID agar.

**Table 3:** BB with confirmation positivity for *C. difficile* by swabs and patients.

| Location | # Swabs | # Patients | Positive: Tox+<br>CD |  | Negative: Tox-<br>CD |  | Negative: No CD |  |
| --- | --- | --- | --- | --- | --- | --- | --- | --- |
|  |  |  | Swabs<br># (%) | Patients<br># (%) | Swabs<br># (%) | Patients<br># (%) | Swabs<br># (%) | Patients<br># (%) |
| BWH (Brigham and Women's Hospital)* | 764 | 509 | 80 (10) | 72 (14) | 28 (4) | 21 (4) | 656 (86) | 479 (94) |
| FH (BWH Faulkner Hospital) | 11 | 10 | 3 (27) | 3 (30) | 1 (9) | 1 (10) | 7 (64) | 7 (70) |
| NWH (Newton Wellesley Hospital) | 22 | 19 | 5 (23) | 4 (21) | 2 (9) | 2 (11) | 15 (68) | 15 (79) |
| **Total** | **797** | **538** | **88 (11)** | **79 (15)** | **31 (4)** | **24 (4)** | **678 (85)** | **501 (93)** |

**Legend:** Results of BB with confirmation testing, showing numbers and percentages, in parentheses, of positive and negative swabs relative to total number of swabs or patients tested. Note: the total percentage of patient samples exceeds 100% due to the longitudinal sampling and variable *C. difficile* status of patients over the surveillance period. Asterix (\*) for BWH indicates BWH ICU and DFCI oncology inpatients undergoing CD surveillance at the BWH Clinical Laboratories.

### Isolate Recovery from BB-positive tubes

For additional strain analyses, we evaluated capacity to extract DNA and cultivate *C. difficile* isolates from 1mL BB aliquots mixed with 0.4 ml sterile 60% glycerol to a final concentration of 17% glycerol, followed by storage at –80°C. All *C*. *difficile* isolates (n=8 strains) were recovered anaerobically on ChromID agar at 7 months of storage, while 88% of isolates (7 of 8 strains tested) were recovered at 12-months of storage. DNA extracted from frozen materials supported PCR detection of GDH and toxin B, while DNA extracted from re-isolated strains supported genome sequencing and analyses (Methods).

### Prospective *C. difficile* surveillance

To scale a *C. difficile* surveillance program, we prospectively tested 797 rectal swabs from ICU and oncology unit inpatients sent to the Clinical Microbiology laboratory at Brigham and Women’s Hospital, a tertiary academic medical center laboratory that also tests cancer inpatients from the Dana-Farber Cancer Institute, and the Clinical Microbiology laboratories at community hospitals Brigham and Women’s Faulkner Hospital (FH) and Newton-Wellesley Hospital (NWH). Surveillance for *C. difficile* colonization evaluated 538 inpatient oncology and ICU patients. Within this cohort, 509 patients originated from BWH and DFCI inpatients, 19 from NWH and 10 from FH. The percent of female patients across sites was 48%, 68%, and 39%, respectively and the mean age and range was 64 at BWH/DFCI (17-99 years), 70 at NWH (33-86 years), and 64 at FH (30-87 years). After completion of VRE surveillance testing, rectal swabs were stored at 4°C for up to 3 days prior to inoculation into BB (Methods) (1). Swabs were inoculated into BB and incubated at 37°C for up to 48h. Among swabs, 69% were resulted by 48h as negative per no color change occurring in BB (Tables 3, 4). Among BB-positive tubes, relative to total swabs tested, 15.8% (126/797) were GDH and Tox A/B negative indicating no *C. difficile* colonization; 2.8% (22/797) were GDH and Tox A/B positive indicating colonization with toxigenic *C. difficile*, and 12.2% (97/797) were GDH positive and Tox A/B negative requiring further confirmatory testing with Xpert. For BB positive tubes proceeding to Xpert confirmation, an additional 8.2% (66/797) tested positive for toxigenic *C. difficile,* and 4.0% (31/797) had toxin-negative *C. difficile* and were resulted as negative (Figure 3, Table 3). The overall positivity rate for toxigenic *C. difficile* was 11% and 4% for non-toxigenic *C. difficile*. The use of CDQCC testing on BB positive tubes produced definitive positive or negative results in 60% of BB positive tubes, requiring Xpert confirmation in 40% of BB positive tubes, or 12% of all surveillance samples (97/797).

**Figure 3.**
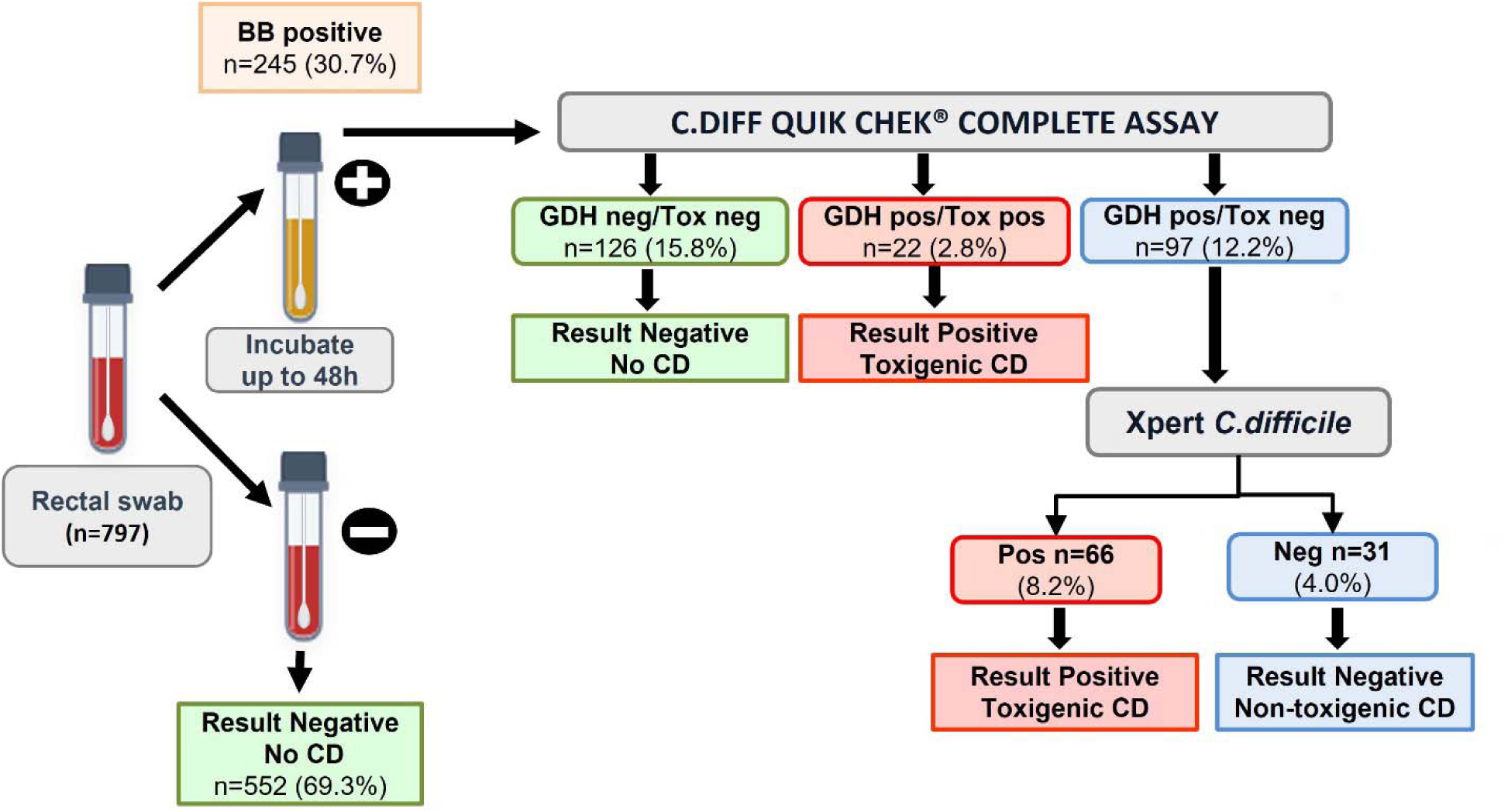
Prospective *C. difficile* surveillance reults using BB with confirmation. **Legend:** This workflow diagram overlays the numbers and percentages that were positive for toxigenic *C. difficile* or negative, where negative samples had no *C. difficile* or non-toxigenic *C. difficile* identified. Rectal swabs were inoculated into BB tubes. Positive BB were tested by Cdiff Quik Chek. Samples GDH positive/Tox negative were further tested by Xpert. The overall positivity for toxigenic CD was 11% and 4% for non-toxigenic CD. This two-step approach allowed definitive results in 88% of all samples, requiring Xpert confirmation in only 12%.

**Table 4:** Time to *C. difficile* results using BB with two-step confirmation.

| Result | 24h | 48h |
| --- | --- | --- |
| Positive: Toxigenic CD | n=28 (3.5%) | n=60 (7.5%) |
| Negative: Non-toxigenic CD | n=11 (1.4%) | n=20 (2.5%) |
| Negative: No CD* | n=23 (2.8%)* | n=655 (82.2%) |

**Legend :** Prospective surveillance results (n=797 swabs) at 24 and 48h of BB with confirmation. Numbers indicate total and overall percentages of the following results: positive for toxigenic *C. difficile*, negative showing non-toxigenic *C. difficile*, and negative showing no *C. difficile* detected samples. Asterix (\*) indicates CD-negative samples identified at 24h from BB+ tubes that were GDH negative by the QuickCheck Cdiff Complete rapid EIA. The remaining "No CD" tubes at 48h showed no color change and were resulted as negative.

Positive results were called to the infection control teams upon confirmation of positive samples, at 24 or by 48h of incubation (Table 4). BB with confirmation identified 32% of patients carrying toxigenic *C. difficile* at 24h and the remaining 68% at 48h. No patients colonized with toxigenic *C. difficile* developed CDI during the 24-48 period for BB-surveillance testing to be resulted.

## Discussion

To scale detection of patient colonization with toxigenic *C. difficile,* we developed a highly sensitive method that leverages *ex vivo* enrichment of *C*.*difficile* in BB. The *ex vivo* biological amplification of *C. difficile* generates sufficient biomass to support robust detection of toxigenic strains with the confirmatory methods. BB-positive tubes were first evaluated with the *C. diff* Quick Check rapid EIA for *C. difficile* GDH and toxins A/B, followed by Xpert PCR detection of toxin gene carriage in samples that were GDH+ but negative for antigenic toxin A or B (Figure 1B), a situation that can occur with populations of *C. difficile* that may not be sufficiently stressed to elaborate toxins (23). The use of BB and both confirmatory assays are off-label and required their validation as lab-developed tests (LDTs). Once implemented, this approach eliminated the need for local anaerobic cultivation while leveraging *C. difficile* test methods already in use for testing stool samples from patients with suspected CDI.

BB with confirmation demonstrated greatly improved sensitivity of detection for *C. difficile* to <5 CFU per swab, a finding consistent with prior studies (24), and detected additional patients colonized with toxigenic *C. difficile* that were missed by ChromID agar. The approach also demonstrated a 100-fold more sensitive threshold for detection of asymptomatic *C. difficile* colonization versus direct swab testing by Xpert, which is optimized for diagnosing patients with the higher pathogen biomass seen in CDI.

BB with confirmation increased our detection of patient colonization on surveilled units to 11% across inpatient and community hospital settings, as compared to rates of 7.4% by ChromID/Xpert (1). The BB method also identified patients colonized only with non-toxigenic *C. difficile*, which occurred in 4% of surveillance samples, and which may provide protection against CDI given use of such strains as live bacteriotherapeutic products (LPBs) for recurrent CDI (25, 26).

BB with confirmation demonstrated 100% sensitivity and 92% specificity for detection of toxigenic *C*.*difficile*. BB’s selective conditions for *C. difficile* culture ruled out the majority of patient samples that required confirmatory testing by EIA or molecular methods, significantly reducing the overall number of EIA and molecular tests needed to support surveillance testing. Recovery of *C*. *difficile* isolates from BB performed well to support additional strain-level analyses Recovered strains supported PCR and genome sequencing.

Our findings demonstrate a simplified workflow that may be scaled to support *C. difficile* surveillance across healthcare settings. Additional EIA and PCR assays, including the BD MAX^TM^ Cdiff test or targeted qPCR, may also be suitable to confirm presence of toxigenic *C*. *difficile* in BB-positive cultures. When integrated within infection prevention programs, identification of patient colonization with toxigenic *C. difficile* can enable specific targeting of interventions to prevent CDI and asymptomatic transmission to naive patients.

## Data Availability

All data produced in the present work are contained in the manuscript.

## Acknowledgements

We thank the Crimson Core at Mass General Brigham for logistical support, Jacinta Foncha at Brigham and Women’s Faulkner Hospital and Beckie Hastings at Newton Wellesley Hospital for clinical microbiologic support. Funding provided by the Hatch Family Foundation (Bry), NIH grants K01AI180357 (Cersosimo), the Harvard Digestive Diseases Center’s P30DK034854, a capital grant from the Massachusetts Life Sciences Center (Bry) and CDC Prevention Epicenter grant U54CK000611 (Klompas).

## Notes

### Competing Interest Statement

The authors have declared no competing interest.

### Clinical Trial

NCT05389904

### Author Declarations

The IRB of Mass General Brigham gave ethical approval for this work under IRB 2005P001742 (Bry, PI) for clinical assay development and validation. The IRB of Mass General Brigham gave ethical approval for this work under IRB 2022P000819 (Baker PI) for prospective surveillance under clinical trial NCT05389904. Rectal swabs submitted for VRE ICU surveillance at Brigham and Women's Hospital (BWH), Brigham and Women's Faulkner Hospital (FH), Newton Wellesley Hospital (NWH), and inpatient oncology units for DFCI were retrieved after completion of VRE surveillance testing for conducting the C. difficile surveillance.

